# Sero-Prevalence of Covid-19 among workers in Malaysia

**DOI:** 10.1101/2022.01.16.22269388

**Authors:** Noorliza Mohamad Noordin, Aziyati binti Omar, Ishmah Hana Isharudin, Riasah Idris, Yukie Chem, Intan Surianne Mat Sahat, Selvanesan Sengol, Zirwatul Adilah Aziz, Zhuo-zhi Lim, Teck-Onn Lim

**Affiliations:** National Public Health Laboratory (NPHL) Malaysia; Healthy Malaysia Society

**Keywords:** Serosurvey, Prevalence, Covid-19, Malaysia

## Abstract

From the beginning of the pandemic in Feb 2020, Malaysia has been through 4 waves of outbreak, the magnitude of each wave is several orders larger than the preceding one. By the end of the fourth wave in October 2021, Malaysia has among the highest death toll in Asia, cumulative incidence of confirmed cases has reached 7.0% (>30% in Klang Valley). However it remains uncertain what is the true proportion of the population infected.

We conducted a sero-survey on 1078 workers from 17 worksites in Klang Valley and Perak between July and September 2021. We tested them for SARS-CoV-2–specific antibodies using Ecotest, a lateral flow immunoassay (LFIA). The ability of antibody testing to detect prior infection depends on the assay and sero-reversion. We therefore adjusted the prevalence estimates to correct for potential misclassification bias due to the use of LFIA and sero-reversion using test sensitivity and specificity results estimated from an independent validation study.

The mean age of the workers was 32 years, 89% were male and migrant workers comprised 81% of all subjects, 59% the subjects were from Klang valley. 33% of workers had prior RT-PCR confirmed Covid-19 infections. We estimated 82.2 percent of workers had been infected by Covid-19 by July-September 2021. Prevalence was 99.9% among migrant workers and 12.1% among local workers. Klang Valley, the most industrialized region in Malaysia where most migrant workers are found, had 100% prevalence, giving an infection-to-case ratio (ICF) of ∼3.

Our sero-prevalence results show that the incidence of Covid19 is extremely high among migrant workers in Malaysia, consistent with findings from other countries such as Kuwait and Singapore which also hosted large number of migrant workers.

## Introduction

Malaysia has emerged from the most recent wave of Covid-19 infection between June and October 2021 with among the highest death toll in Asia. By October 2021, just before the national lockdown was lifted, the cumulative incidence of confirmed Covid-19 had reached 75,550 per million population or 7.5% of the population [1] (Figures 1 and 2).

**Figure 1:**
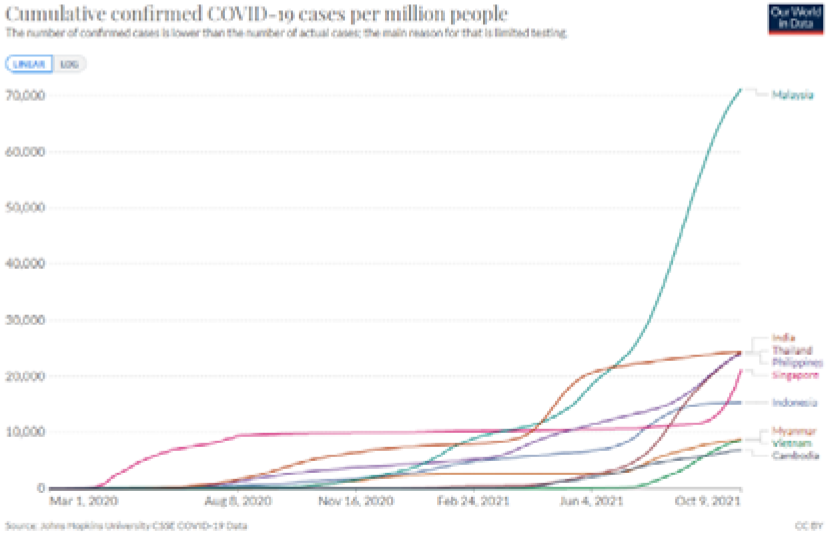
Cumulative confirmed Covid-19 cases per million people in ASEAN & India

**Figure 2:**
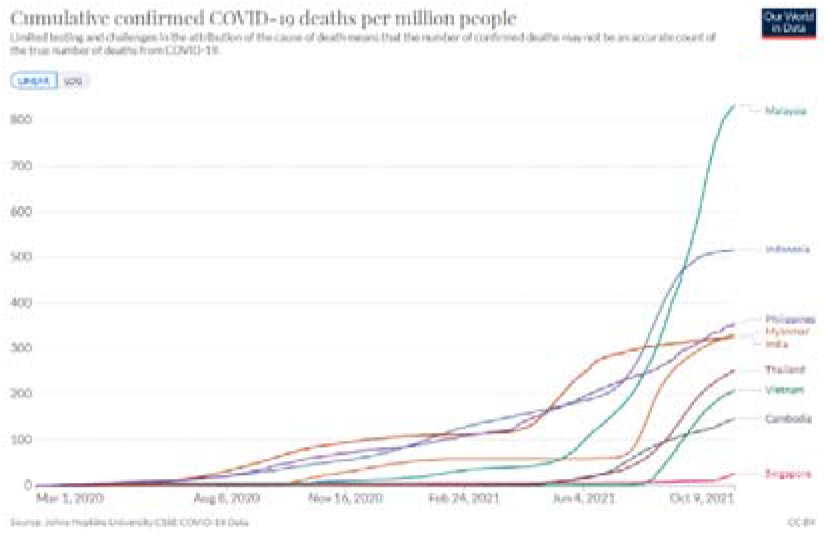
Cumulative confirmed Covid-19 deaths per million people in ASEAN & India

Covid-19 infection is often asymptomatic, as much as 40% of confirmed Covi19 according to one systematic review [2], and a large sero-survey in neighbouring Singapore showed 78% of infected migrant workers had sub-clinical infection [3]. Hence, antibody testing in sero-prevalence survey is necessary to provide information on past exposure to severe acute respiratory syndrome coronavirus 2 (SARS-CoV-2). This is useful for estimating the true burden of Covid-19, and for identifying at-risk groups, such as healthcare workers, migrant workers and nursing home residents. To date, three sero-prevalence studies [4,5,6] have been conducted in Malaysia, as summarized in Table 1. These surveys have found remarkably low prevalence perhaps because they were conducted in 2020 early in the course of the Covid-19 epidemic in Malaysia.

**Table 1:** Data on Sero-prevalence in Malaysia.

| # | Study | Antibody assay | Study Calendar time, location & population | Prevalence estimate |
| --- | --- | --- | --- | --- |
| 1. | COVID-19 antibody surveillance among healthcare workers in a non-COVID designated cardiology centre [4]. | Lateral flow immunoassay | March 2020<br>310 healthcare workers in a single hospital in Sarawak | Age standardized 4.5% |
| 2. | Low post pandemic wave SARS-CoV-2 sero-prevalence in Kuala Lumpur and Selangor, Malaysia [5] | Neutralizing antibody (cPass) | January-June 2020<br>588 stored serum samples at a teaching hospital serving Kuala Lumpur and Selangor | 0.4% |
| 3. | Serology surveillance of SARS-CoV-2 antibodies among healthcare workers in COVID-19 designated facilities in Malaysia [6] | Neutralizing antibody (cPass) | April- May 2020<br>400 healthcare workers in public hospitals in Kuala Lumpur | 0.0% |

An important community risk factor driving Covid-19 infection in Malaysia is the presence of large number of migrant workers, which comprise 22% of our labour force (3.5 million [7] out of 15.7 million in 2020 [8]). This is attested by numerous press reports on frequent occurrences of Covid-19 outbreaks at workplaces [9]. Singapore, a neighboring country, which also hosted large number of migrant workers, has reported infection rate of 56% among its workers in dormitories [10]. And Thailand, another neighboring country hosting large number of migrant workers, similarly attributed a recent surge in Covid-19 to new clusters at the country’s largest shrimp market where numerous migrant workers are employed [11].

In this study, we provide estimates of the sero-prevalence of Covid-19 infection between June-September 2021 in Malaysia, and compare the prevalence between migrant and local workers.

## Methods

The study sample for this sero-survey were subjects who participated in ongoing studies on worksite-based screening and vaccination between July and September 2021. They were recruited from 17 worksites, comprising 17 factories and 2 farms located in Federal Territory and Selangor (jointly named Klang valley) and Perak. The Ministry of Health’s (MOH) Medical and Research Ethics Committee approved the study and all subjects gave informed consent.

These subjects had baseline SARS-CoV-2–specific antibodies tested as part of the studies. They were tested using Ecotest, a lateral flow immunoassay (LFIA). The LFIA test was performed according to the manufacturer’s instructions and was conducted by trained nurses on finger-stick capillary samples taken from the participants.

The ability of antibody testing to detect prior infection depends on the assay, disease severity and timing of testing [12]. LFIA has lower sensitivity and specificity compared to lab-based ELISA test. We therefore validated the test performance of LFIA by repeat testing for a subset of 468 subjects (393 tested negative on LFIA, 75 tested positive) who also had neutralizing antibody assay (cPass GenScript Biotech). Using the neutralizing antibody assay as reference and 20% neutralization inhibition as cutoff, we determined the LFIA has a test sensitivity of 83% and specificity 96%. Antibody levels will decay over time, and antibody response may be transient in mild or asymptomatic infection [13]. Sero-reversion may have occurred among some subjects; that is their antibody level may have dropped below the threshold for positivity at the time of entry into this study. We also determined the prior RT-PCR confirmed Covid-19 status for a subset of 468 subjects. Using their prior RT-PCR test results as reference, we determined that antibody test at study entry has a sensitivity of 28%. We use these estimates of test sensitivity and specificity to adjust the prevalence estimates of Covid-19, in order to correct for potential misclassification bias due to the use of LFIA and sero-reversion [14,15].

## Results

A total of 1078 workers participated in the sero-survey between July and September 2021. Table 1 shows the characteristics of all the participants. The mean age of the workers was 32 years, 89% were male and migrant workers comprises 81% of all subjects, 59% the subjects were from Klang valley. 33% of workers had prior RT-PCR confirmed Covid-19 infections.

We estimated 82.2 percent of workers had been infected by Covid-19 by June-September 2021. Prevalence was 99.9% among migrant workers and 12.1% among local workers. Klang Valley, the most industrialized area in Malaysia where most migrant workers are employed, had 100% prevalence while in Perak 55% of migrant workers had prior exposure.

**Table 1:** Characteristics of workers who participated in Covid-19 sero-survey, Malaysia July-September 2021.

|  | **Local workers** | **Migrant workers** |
| --- | --- | --- |
| Total number | 204 | 874 |
| **Demographic, No. (%)** |  |  |
| Age, year |  |  |
| Mean (SD) | 32 (12) | 32 (7) |
| Median (IQR) | 28 (23,40) | 32 (27,38) |
| Gender, No. (%) |  |  |
| Male | 136 (67%) | 823 (94%) |
| Female | 68 (33%) | 51 (6%) |
| Ethnicity, No. (%) |  |  |
| Malay | 77 (38%) | NA |
| Chinese | 120 (59%) | NA |
| Indian | 7 (3%) | NA |
| Location, No. (%) |  |  |
| Federal Territory & Selangor (Klang valley) | 129 (63%) | 510 (58%) |
| Perak | 75 (34%) | 364 (41%) |
| **Prior RT-PCR confirmed Covid-19** |  |  |
| Total number | 75 | 393 |
| No. (%) positive | 0 (0) | 198 (50%) |

Abbreviation: NA Not applicable

**Table 2:** Sero-prevalence estimates among workers in Malaysia, July to September 2021.

|  | **Local workers** | **Migrant workers** |
| --- | --- | --- |
| Total number | 204 | 874 |
| **Unadjusted estimate, % (95% CI)** |  |  |
| Overall | 4.4%<br>(2.3, 8.3) | 23.8%<br>(21.1, 26.7) |
| Klang valley<br>(Federal Territory and Selangor) | 4.6%<br>(2.1, 10.0) | 30.8%<br>(26.9, 34.9) |
| Perak | 4.0%<br>(1.3, 11.7) | 14.0%<br>(10.7, 17.9) |
| **Estimate adjusted for Sero-reversion and test performance, % (95% CI)** |  |  |
| Overall | 12.1%<br>(8.0, 16.2) | 99.9%<br>(96.7, 100) |
| Klang valley<br>(Federal Territory and Selangor) | 13.0% | 100%<br>(-) |
| Perak | 10.3% | 55.0% |

## Discussion

From the beginning of the pandemic in Feb 2020, Malaysia has been through 4 waves of outbreak, the magnitude of each wave is several orders larger than the preceding one. By the end of the fourth wave in October 2021, we have among the highest death toll in Asia [1], cumulative incidence of confirmed cases has reached 7.0% [1] (>30% in Klang Valley) [16]. Consistent with this, we reported here a sero-prevalence of 99% among migrant workers, 100% in Klang Valley, giving an infection-to-case ratio (ICF) of ∼3 in contrast to the ICF in India of 56.2 [17].

Our result is also consistent with findings from similar countries hosting large number of migrant workers, though more extreme. Kuwait, a Gulf country hosting large number of migrant workers who accounted for >60% of confirmed cases, reported a sero-prevalence of 38.1% in May-June 2020 [18]. In neighbouring Singapore, migrant workers who lived in dormitory accounted for 90% of infections there [19], and a large study in December 2020 found a prevalence of 56% [3], no doubt higher by now.

Malaysia has long been warned that its large number of migrant workers, many of whom are undocumented illegals, working and housed in crowded unhygienic condition, will pose a serious risk of recurrent outbreaks [20,21]. Migrant workers are mostly young healthy adults, a very high percentage of them have asymptomatic infections, thus becoming an important source of transmission in the community [22,23]. Our results show that this has come to pass.

And yet, we have turned a blind eye to this, unable to formulate effective policy to address the risk, simply because there are too many vested interests involved. The Malaysian economy, especially the manufacturing, construction, agriculture and catering sectors, has become heavily dependent on low cost migrant labour to stay afloat. The supply of migrant labour to private employers has itself become a huge industry, and a highly lucrative source of income to politicians and government officers who control issuance of the labour import license, while street bureaucrats like police and immigration officers preyed on employers and migrant workers for bribes. Under such circumstances, it is not surprising that public health interest will take a back seat.

This study has limitations.

- We have used a convenient sample which is not representative of the workers’ population. Given the nature of the migrant labour industry where illegal practices are widespread, there does not exist a national sampling frame to provide a random sample for the study. Indeed just recruiting 17 worksites to agree to participate was challenging. Participating worksites are therefore not likely to be representative of all worksites, they are likely to be more compliant in their labour practices and yet we found such high prevalence of Covid19.
- We have used LFIA to detect the presence of antibody. We have adjusted accordingly for the test imperfect sensitivity and specificity. We have also conducted an independent validation study to estimate the test sensitivity and specificity (rather than rely on manufacturer’s reported results). The validation sample was the same as the subjects (workers) in this study, rather than another study population such as hospitalized patents.
- By July-September 2021, we are 20 month into the pandemic in Malaysia. Many workers are likely to have been infected earlier in the course of the epidemic and will likely have sero-reverted at the time of this study. Indeed we found a positive neutralizing antibody test has a sensitivity of only 28to detect prior exposure among subjects who previously had PCR confirmed Covid19. We have adjusted the estimate accordingly for sero-reversion.

## Data Availability

All data produced in the present study are available upon reasonable request to the authors

